# The current state of COVID-19 in Australia: importation and spread

**DOI:** 10.1101/2020.03.25.20043877

**Authors:** Jessica Liebig, Raja Jurdak, Ahmad El Shoghri, Dean Paini

**Affiliations:** Data61, Commonwealth Scientific and Industrial Research Organisation Brisbane, Queensland, Australia; School of Electrical Engineering and Computer Science, Queensland University of Technology Brisbane, Queensland, Australia; School of Computer Science and Engineering, University of New South Wales Sydney, New South Wales, Australia; Health and Biosecurity, Commonwealth Scientific and Industrial Research Organisation Canberra, Australian Capital Territory, Australia

## Abstract

**Background:** The rapid global spread of coronavirus disease (COVID-19) is unprecedented. The outbreak has quickly spread to more than 100 countries reporting over 100,000 confirmed cases. Australia reported its first case of COVID-19 on 25^th^ January 2020 and has since implemented travel restrictions to stop further introduction of the virus.

**Methods:** We analysed daily global COVID-19 data published by the World Health Organisation to investigate the spread of the virus thus far. To assess the current risk of COVID-19 importation and local spread in Australia we predict international passenger flows into Australia during 2020.

**Findings:** Our analysis of global data shows that Australia can expect a similar growth rate of reported cases as observed in France and the United States. We identify travel patterns of Australian citizens/residents and foreign travellers that can inform the implementation of new and the alteration of existing travel restrictions related to COVID-19.

**Interpretation:** Our findings identify the risk reduction potential of current travel bans, based on the proportion of returning travellers to Australia that are residents or visitors. The similarity of the exponential growth in the epidemic curve in Australia to other countries guides forecasts of COVID-19 growth in Australia, and opportunities for drawing lessons from other countries with more advanced outbreaks.

## Introduction

Coronavirus disease (COVID-19) is caused by severe acute respiratory syndrome coronavirus 2 (SARS-CoV-2), a novel virus that has not previously been observed in humans [1]. Since its introduction into the human population in late 2019, the virus has caused a global outbreak that prompted the World Health Organisation (WHO) to declare a public health emergency of international concern [2]. On 8^th^ March 2020, the global number of confirmed cases, reported by more than 100 countries, rose above 100,000 [3]. Three days later the WHO announced COVID-19 to be a pandemic [4]. Global air travel has likely been one of the major drivers of the rapid long-distance spread [5].

COVID-19 is a zoonotic disease and was likely introduced from bats into the human population [6]. The first cluster of cases detected at the end of 2019 has been linked to a seafood marked in Wuhan, China [6]. On 13^th^ January, Thailand reported the first imported case of COVID-19. Two days later a second imported case was detected in Japan. The third country outside China to report an im-ported case of COVID-19 was South Korea [7]. Figure 1 shows the number of countries that have reported cases of COVID-19 since the beginning of the outbreak together with a timeline of important events. The graph shows that after an initial growth between 10^th^ January and 1^st^ February 2020 the number of countries reporting cases of COVID-19 remained steady until 24^th^ February. There has since been a steady increase in the number of reporting countries, with an average of six additional countries reporting COVID-19 cases each day.

**Figure 1:**
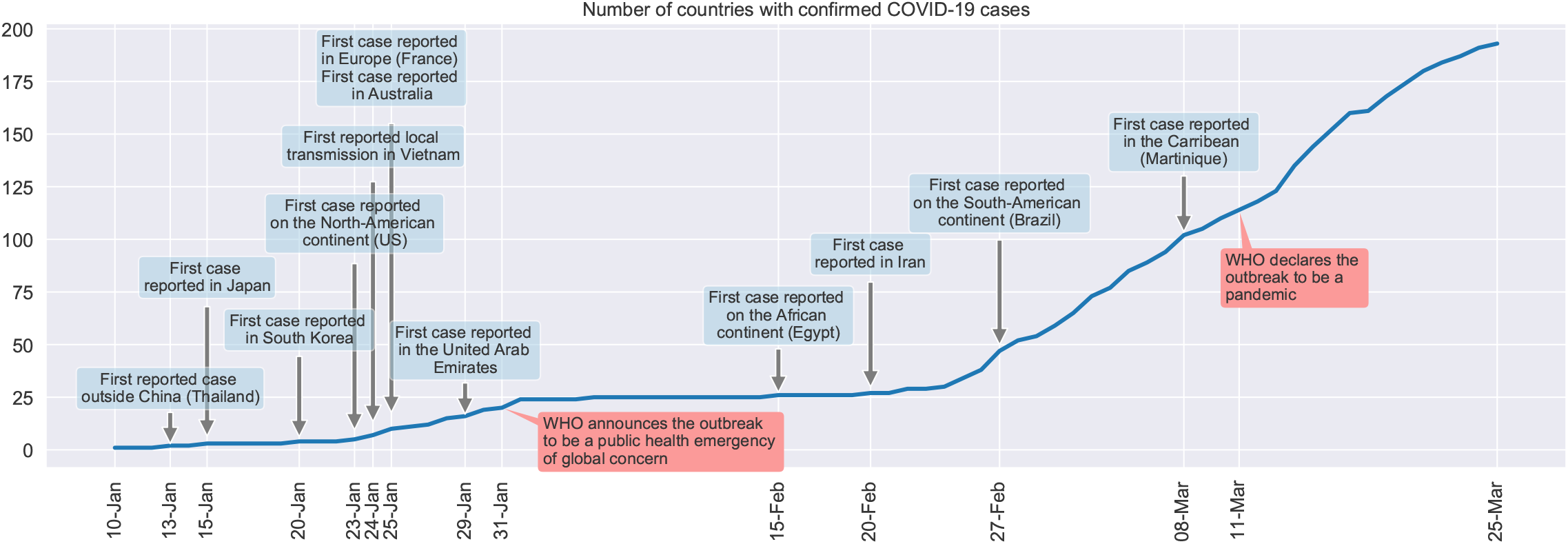
The cumulative number of countries that have reported COVID-19 cases over time.

Initially most reported cases outside China showed a travel history to Wuhan. The first locally acquired case of COVID-19 was reported on 24^th^ January in Vietnam, suggesting the possibility of human-to-human transmission. It is now evident that humans can transmit the disease to each other, mainly through droplets and via surfaces that allow the virus to survive [8, 9, 10]. As of 25^th^ March, among the 193 countries, territories and areas that have confirmed cases of COVID-19, 125 (64.77%) have reported local transmission [11].

In Australia the first imported case was reported on 25^th^ January 2020 in New South Wales and began showing symptoms of the disease on 13^th^ January. In the week following the first case report, another eleven cases of COVID-19 were confirmed. Investigations revealed that all twelve individuals acquired the virus in China [12]. During the week ending 8th February, the state of Queensland reported limited local transmission of the virus [13]. A previously imported case from China, who was part of a travel group, infected three group members prior to being isolated. Further transmission was prevented by quarantining the travel group. The three secondary cases began to show symptoms during the quarantine period. There is now further evidence of local transmission inside Australia, with at least 406 cases being local transmissions [14]. As at 3:00 pm on 25th March 2020, Australia has reported a total of 2,423 COVID-2019 cases across all of its states and territories [15].

In response to the rising number of COVID-19 cases globally and in Australia, the Australian government began to implement travel restrictions on 1^st^ February, denying entry to travellers who had stayed or transited through mainland China in the past 14 days [16]. While Australian citizens, permanent residents, immediate family members of citizens and permanent residents, New Zealand citizens residing in Australia and diplomats are exempt from the restrictions, they are required to self-isolate at home for 14 days upon their return. On 1^th^ March the travel restrictions were extended to individuals travelling from Iran. Travellers from South Korea and Italy were included on 5^th^ March and 11^th^ March, respectively. On 15^th^ March the Australian government announced that every individual who enters Australia is required to self-isolate for 14 days, regardless of the country of origin. The travel restrictions were further tightened on 20^th^ March, with only Australian citizens, permanent residents and immediate family members being allowed to enter Australia.

## Methods

### Epidemic curves

To illustrate the progression of the COVID-19 pandemic in different countries, we plot the cumulative epidemic curves by reporting date. In order to compare the growth of cases across different countries, we align the curves relative to the date when a country reached 100 reported cases. To test for exponential growth, we plot the epidemic curves on semilogarithmic axes. That is, if the growth of reported cases is exponential, the cumulative epidemic curve on a semilogarithmic plot will form a straight line.

### Fitting exponential curves

To predict the expected number of reported cases over the coming days in Australia, we fit an exponential curve to the observed reported cases by minimizing the sum of the squared residuals.

### Predicting travel patterns

We predict the number of arrivals into Australia for the year 2020 based on ten years of historical data (January 2010 - December 2019) using a seasonal autoregressive integrated moving average (SARIMA) model, denoted ARIMA(*p, d, q*)(*P, D, Q*)_*m*_, where *p* is the autoregressive order of the non-seasonal part, *d* is the order of differencing that is required to remove the trend from the time series, *q* is the non-seasonal order of the moving-average model and *m* is the number of observations per year. Parameters *P, D* and *Q* are the seasonal counterparts of *p, d* and *q*. The autoregressive part of the model uses a linear combination of previous values of the time-series. That is, the autoregressive order *p* indicates the number of previous values considered in the prediction. Table 1 lists the input parameters of the SARIMA models used to predict passenger flows from different countries.

**Table 1:** Model parameters used for the different SARIMA models. Parameters *p, d* and *q* are non-seasonal and denote the autoregressive order, the degree of differencing and the order of the moving average, respectively. Parameters *P, D, Q* are the seasonal counterparts and *m* denotes the number of observations per seasonal cycle.

| Country | Traveller | $p$ | $d$ | $q$ | $P$ | $D$ | $Q$ | $m$ |
| --- | --- | --- | --- | --- | --- | --- | --- | --- |
| China | Residents | 2 | 0 | 2 | 0 | 1 | 2 | 12 |
| China | Visitors | 1 | 1 | 1 | 1 | 1 | 2 | 12 |
| France | Residents | 0 | 1 | 1 | 0 | 1 | 2 | 12 |
| France | Visitors | 1 | 0 | 1 | 1 | 1 | 1 | 12 |
| Germany | Residents | 0 | 0 | 0 | 1 | 1 | 2 | 12 |
| Germany | Visitors | 1 | 0 | 1 | 0 | 1 | 1 | 12 |
| India | Residents | 0 | 0 | 3 | 0 | 1 | 1 | 12 |
| India | Visitors | 0 | 0 | 3 | 0 | 1 | 1 | 12 |
| Indonesia | Residents | 1 | 0 | 1 | 0 | 1 | 1 | 12 |
| Indonesia | Visitors | 1 | 0 | 0 | 0 | 1 | 0 | 12 |
| Iran | Residents | 0 | 1 | 1 | 0 | 1 | 1 | 12 |
| Iran | Visitors | 1 | 0 | 2 | 0 | 1 | 1 | 12 |
| Italy | Residents | 1 | 0 | 0 | 1 | 1 | 2 | 12 |
| Italy | Visitors | 2 | 1 | 3 | 1 | 1 | 1 | 12 |
| Japan | Residents | 1 | 0 | 1 | 0 | 1 | 1 | 12 |
| Japan | Visitors | 1 | 1 | 1 | 0 | 1 | 1 | 12 |
| Malaysia | Residents | 1 | 0 | 1 | 0 | 1 | 1 | 12 |
| Malaysia | Visitors | 0 | 0 | 3 | 0 | 1 | 1 | 12 |
| New Zealand | Residents | 0 | 0 | 2 | 0 | 1 | 1 | 12 |
| New Zealand | Visitors | 0 | 0 | 0 | 0 | 1 | 1 | 12 |
| Singapore | Residents | 3 | 0 | 0 | 0 | 1 | 1 | 12 |
| Singapore | Visitors | 2 | 0 | 3 | 0 | 1 | 1 | 12 |
| South Korea | Residents | 0 | 0 | 0 | 0 | 1 | 1 | 12 |
| South Korea | Visitors | 1 | 0 | 2 | 0 | 1 | 0 | 12 |
| Spain | Residents | 1 | 0 | 0 | 0 | 1 | 1 | 12 |
| Spain | Visitors | 1 | 0 | 1 | 0 | 1 | 1 | 12 |
| Thailand | Residents | 0 | 1 | 1 | 0 | 1 | 2 | 12 |
| Thailand | Visitors | 1 | 0 | 0 | 1 | 1 | 0 | 12 |
| United Kingdom | Residents | 0 | 0 | 2 | 0 | 1 | 1 | 12 |
| United Kingdom | Visitors | 0 | 0 | 1 | 0 | 1 | 1 | 12 |
| United States | Residents | 0 | 1 | 1 | 1 | 1 | 0 | 12 |
| United States | Visitors | 1 | 0 | 1 | 1 | 1 | 1 | 12 |

Before finding the parameters for the SARIMA model, we transformed the individual time-series using a Box-Cox transformation to give the data a normal shape [17]. The transformed time-series was tested whether it is stationary using the Kwiatkowski-Phillips-Schmidt-Shin test [18]. To find the model with the best fit we perform a step-wise search over the model space [19] and chose the model with the lowest AIC (Akaike’s Information Criterion). We considered the following parameter ranges: *p* ∈ [0, 5], *d* ∈ [0, 2], *q* ∈ [0, 5], *P* ∈ [0, 2], *Q* ∈ [0, 2]. All calculations have been carried out with Python’s pmdarima statistical library.

### Data

We extracted the number of reported COVID19 cases from the daily situation reports published by the WHO. All reports are available at https://www.who.int/emergencies/diseases/novel-coronavirus-2019/situation-reports.

Australia is one of the few countries that does not have any land borders and, in addition, has strict border controls in place. Every individual who enters Australia is required to fill in a passenger arrival card that records information about the traveller as well as the location and length of the overseas stay. The data collected through arrival cards is publicly available from the Australian Bureau of Statistics in aggregate form (https://data.gov.au/dataset/ds-dga-5a0ab398-c897-4ae3-986d-f94452a165d7/details).

## Results

### Growth of reported cases in Australia

The data published by the WHO shows that the time between the first reported case in a country and the number of cases in that country rising above 20 is on average 11 days. An epidemic is unlikely to fade out without intervention after 20 individuals have been infected and will begin to follow an exponential growth [20]. In Figure 2 we have overlaid the epidemic curves of Australia and the nine countries that have reported the highest number of cases. For comparison, each epidemic curve is plotted from the time the reported number of cases rose above 100 on semi-logarithmic axes. The epidemic curves of the European countries, the United States and Australia closely follow straight lines, suggesting exponential growth rates. The Chinese, Iranian and South Korean curves show an initial exponential growth, for approximately seven days, before the reported number of cases increases less rapidly.

**Figure 2:**
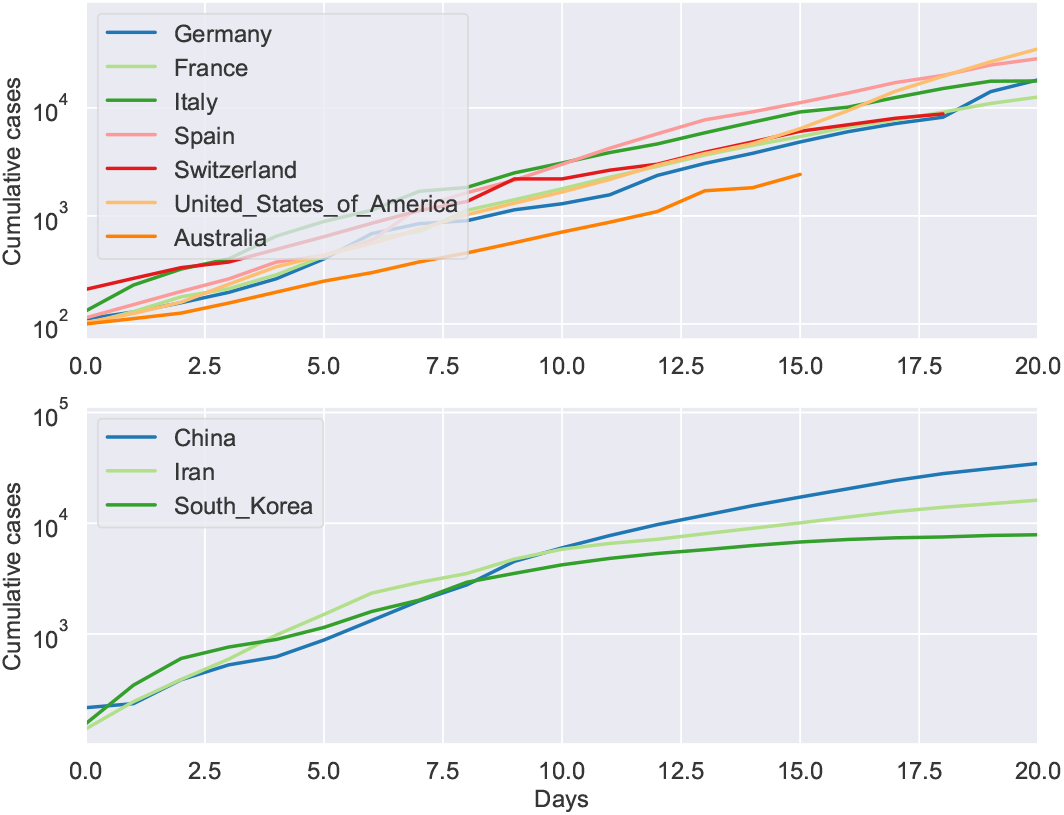
The cumulative epidemic curves of Australia and the nine countries with the highest number of COVID-19 cases. The curves are aligned on the day where the individual countries reached 100 reported cases.

Australia’s cumulative epidemic curve follows that of Germany most closely. Given this observation and the fact that the reported number of COVID-19 cases in Australia has risen well above 20, the risk of a continuing exponential increase in the number of cases is substantial.

Figure 3 shows the cumulative number of reported cases in Australia through to 25^th^ March 2020 (left panel). After 29^th^ February the number of reported cases increases linearly on the semi-logarithmic plot, indicating an exponential increase in cases. The right panel of Figure 3 shows the cumulative number of cases reported in each of the Australian states and territories. The curves of New South Wales, Victoria and Queensland indicate exponential growth.

**Figure 3:**
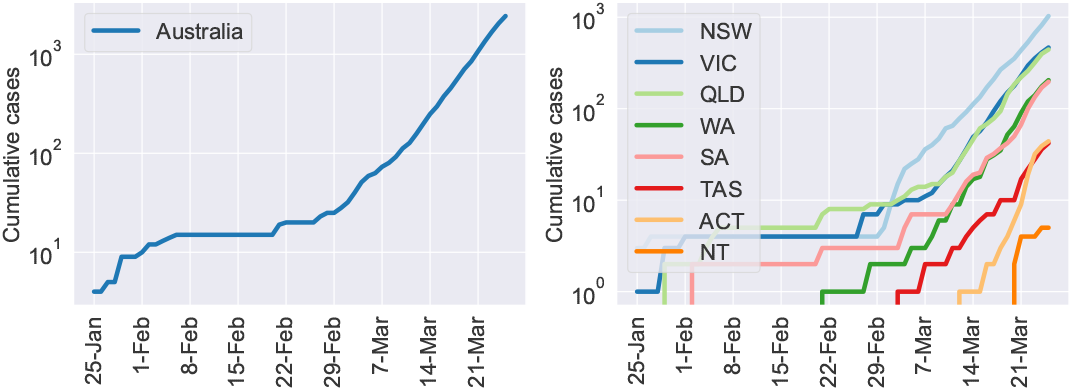
The cumulative number of COVID-19 cases in Australia and in each of its states.

Based on the observations made in other countries around the world that have reported a high number of COVID-19 cases, it is expected that numbers in Australia will continue to rise exponentially. We estimate the number of cases expected to be reported during the next seven days, until 1^st^ April, by fitting the observed data to an exponential function. Figure 4 shows the cumulative number and predicted cumulative number of cases together with the 95% confidence interval (shaded area). If the number of COVID-19 cases continues to rise exponentially, in a similar manner as observed in previous weeks, we predict that by 1^st^ April the number of reported COVID-19 cases in Australia could be as high as 11,908.

**Figure 4:**
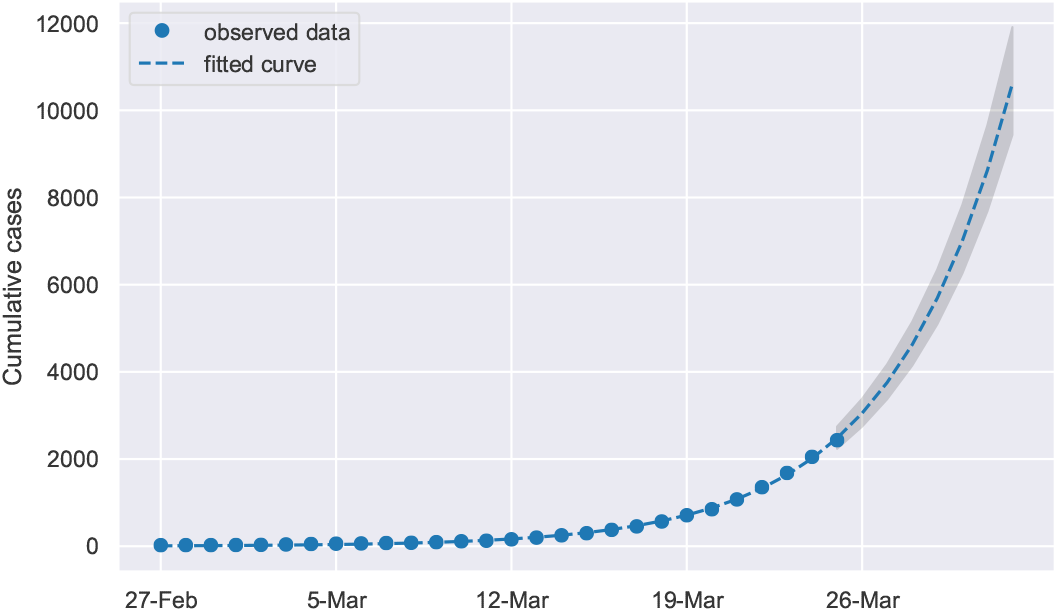
Predicted cumulative number of COVID-19 cases in Australia. We predicted the number of COVID-19 cases in Australia until 1^st^ April (dashed line) by fitting an exponential function to the cases that have been reported so far (blue circles). The equation of the fitted curve is *y* = 9.0822*e*^0.2078*x*^. The shaded area corresponds to the 95% confidence interval of the predictions.

### Travel patterns

Although travel restrictions can be effective in the intervention of infectious disease spread, they are temporary and difficult to sustain in the long term [21]. In the case of travel bans being lifted, a rapid increase in the number of imported cases can be expected.

In 2019, air passengers from New Zealand formed the largest group (≈ 2, 622, 967) of arrivals into Australia, followed by arrivals from China (≈ 1, 999, 857) and the United States (≈ 1, 700, 860). Table 2 lists the ten countries with the most arrivals into Australia in 2019.

**Table 2:** The ten countries with the most arrivals into Australia in 2019.

| Country | Arrivals in 2019 |
| --- | --- |
| New Zealand | 2,884,668 |
| China | 2,150,628 |
| United States | 1,868,148 |
| Indonesia | 1,636,718 |
| United Kingdom | 1,429,122 |
| Japan | 1,030,027 |
| India | 936,134 |
| Singapore | 901,200 |
| Malaysia | 676,811 |
| Thailand | 657,844 |

We predict the number of arrivals into Australia for the year 2020 based on ten years of historical data (January 2010 December 2019) using a seasonal autoregressive integrated moving average (SARIMA) model. Arrivals are divided into two groups, Australian citizens/residents and visitors. The predicted passenger flows for 2020 are shown in Figure 5. The light blue and dark blue bars are our predictions of the number of arriving citizens/residents and visitors, respectively, for the year 2020. The error bars indicate the 95% confidence intervals of the predictions. Black circles show the number of arrivals during 2019.

**Figure 5:**
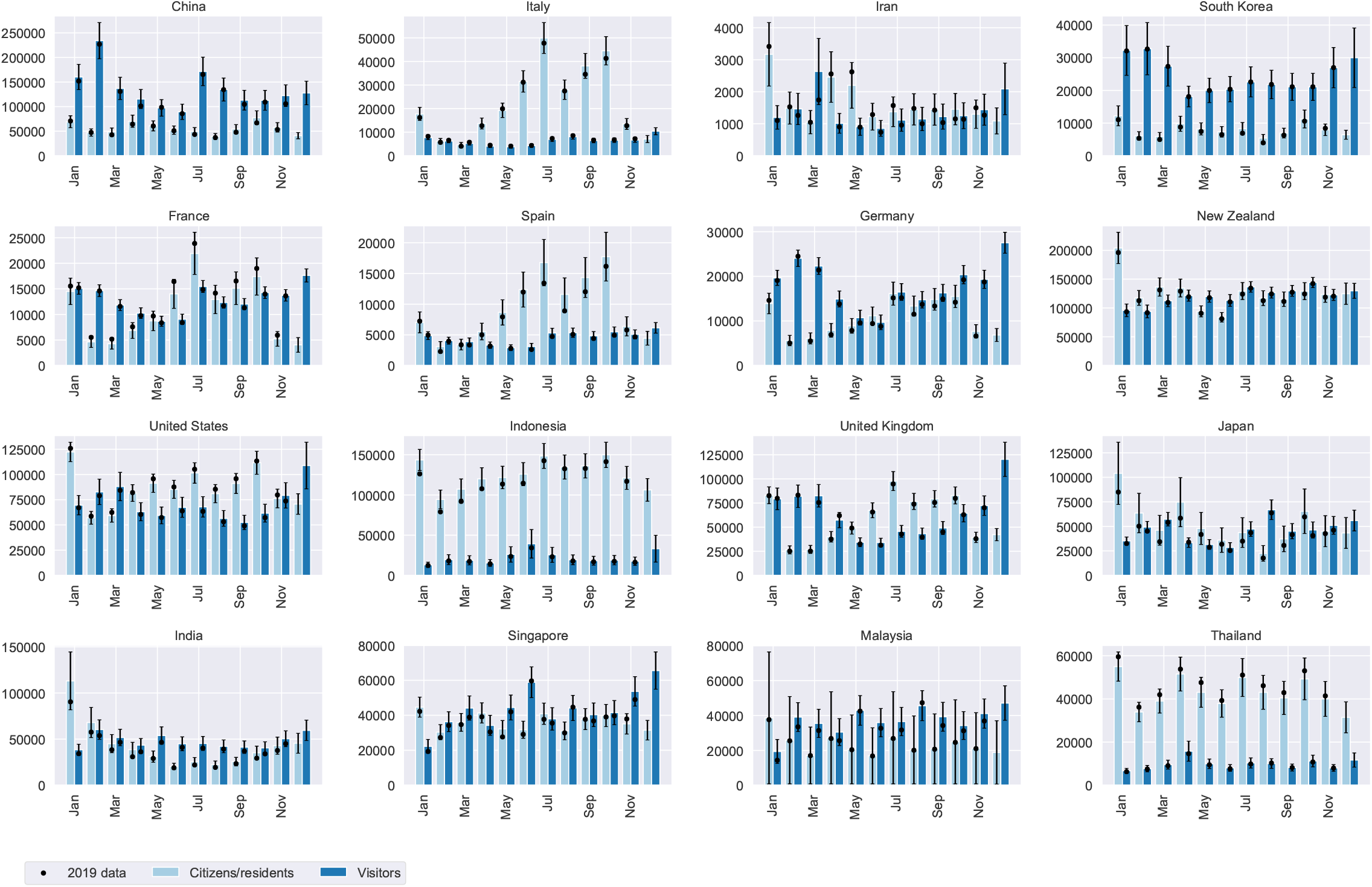
Predicted arrivals of Australian citizens/residents and visitors during 2020. We used ten years of Australian arrival card data to predict arrivals of Australian citizens/residents and visitors into the country. The light blue bars show predictions for citizens/residents and the dark blue bars show predictions for visitors. The error bars indicate the 95% confidence interval of the predictions and black circles show the monthly number of arrivals during 2019.

Among the countries with the largest number of reported COVID-19 cases, China and South Korea stand out with the number of visitors far exceeding the number of returning citizens/residents throughout the year. Australian citizens and residents clearly dominate return travel from Italy and Spain over the northern hemisphere’s summer. The same pattern is observed to a lesser extend for arrivals from France and the United Kingdom. Countries, such as Indonesia and Thailand, have a high ratio of citizens over visitor among travellers into the country.

## Discussion

Our results indicate that without further intervention, Australia can expect a similar growth in the number of reported cases as observed in Germany. We predict that the number of cases in Australia could rise from 2,423 to 11,908 within one week if no intervention takes place.

Our predictions of travel patterns can help policy makers weigh the COVID-19 risk posed by travellers from different countries, as well as the potential reduction in this risk by implementing travel bans and quarantine rules that only apply to foreign travellers. The risk reduction in COVID-19 importations into Australia as a result of travel bans varies across the different source countries. The risk reduction when travel bans are applied to countries where traveller volumes into Australia are dominated by visitors, such as China and South Korea, can be substantial. In contrast, the risk reduction for travel bans relating to countries with a dominant number of returning Australian citizens or residents, such as Italy, Spain, Thailand, and Indonesia, will have a smaller impact on COVID-19 importations, as the citizens and residents can still enter the country.

The travel patterns also reveal seasonal trends in traveller volumes into Australia that vary across countries. Travellers from China, for instance, who are mostly visitors, peak in February and July, aligning with the start of the two main semesters in Australian universities. There are more than 100,000 Chinese students studying at Australian universities. The three Mediterranean European countries, Italy, Spain, and France, have clear peaks in incoming travellers to Australia around the northern hemisphere summer, where Australian citizens and travellers return in significantly larger numbers. Some countries where returning residents dominate incoming travel, such as Japan and Thailand, demonstrate spikes in returning travellers around January, April, July, and October, coinciding with school holidays in Australia. Such seasonal patterns can help authorities plan for travel policies that consider the varying risk of COVID-19 importations into Australia over time from different countries.

## Data Availability

All datasets referred to in the manuscript is publicly available.

https://data.gov.au/dataset/ds-dga-5a0ab398-c897-4ae3-986d-f94452a165d7/details

https://www.who.int/emergencies/diseases/novel-coronavirus-2019/situation-reports

https://www.covid19data.com.au/

## Contributors

JL conceived the study and performed the analysis. AES wrote the code to clean and process the data. RJ and DP assisted with the analysis and contributed to the interpretation of the results. All authors edited and approved the final manuscript.

## Notes

### Competing Interest Statement

The authors have declared no competing interest.

### Funding Statement

No funding source

## References

[1] Chih-Cheng Lai, Tzu-Ping Shih, Wen-Chien Ko, et al. Severe acute respiratory syndrome coronavirus 2 (SARS-CoV-2) and coronavirus disease-2019 (COVID-19): The epidemic and the challenges. Int. J. Antimicrob. Agents, 55:105924, 2020.

[2] World Health Organisation. Novel Coronavirus (2019-nCoV) Situation Report - 11, 2020. https://www.who.int/docs/default-source/coronaviruse/situation-reports/20200131-sitrep-11-ncov.pdf?sfvrsn=de7c0f7_4 (accessed: Feb 01, 2020).

[3] World Health Organisation. Coronavirus disease 2019 (COVID-19) Situation Report – 48, 2020. https://www.who.int/docs/default-source/coronaviruse/situation-reports/20200308-sitrep-48-covid-19.pdf?sfvrsn=16f7ccef_4 (accessed: Mar 09, 2020).

[4] World Health Organisation. Coronavirus disease 2019 (COVID-19) Situation Report – 51, 2020. https://www.who.int/docs/default-source/coronaviruse/situation-reports/20200311-sitrep-51-covid-19.pdf?sfvrsn=1ba62e57_10 (accessed: Mar 12, 2020).

[5] Cristian Biscayart, Patricia Angeleri, Susana Lloveras, et al. The next big threat to global health? 2019 novel coronavirus (2019-nCoV): What advice can we give to travellers?-Interim recommendations January 2020, from the Latin-American society for Travel Medicine (SLAMVI). Travel Med. Infect. Dis., 33:101567, 2020.

[6] Peng Zhou, Xing-Lou Yang, Xian-Guang Wang, et al. A pneumonia outbreak associated with a new coronavirus of probable bat origin. Nature, 579:1–4, 2020.

[7] World Health Organisation. Novel Coronavirus (2019-nCoV) Situation Report - 1, 2020. https://www.who.int/docs/default-source/coronaviruse/situation-reports/20200121-sitrep-1-2019-ncov.pdf?sfvrsn=20a99c10_4 (accessed: Mar 16, 2020).

[8] Jasper Fuk Woo Chan, Shuofeng Yuan, Kin Hang Kok, et al. A familial cluster of pneumonia associated with the 2019 novel coronavirus indicating person-to-person transmission: a study of a family cluster. Lancet, 2020.

[9] Isaac Ghinai, Tristan D McPherson, Jennifer C Hunter, et al. First known person-to-person transmission of severe acute respiratory syndrome coronavirus 2 (SARS-CoV-2) in the USA. Lancet, 2020.

[10] Rachael Pung, Calvin J Chiew, Barnaby E Young, et al. Articles Investigation of three clusters of COVID-19 in Singapore: implications for surveillance and response measures. Lancet, 2020.

[11] World Health Organisation. Coronavirus disease 2019 (COVID-19) Situation Report - 65, 2020. https://www.who.int/docs/default-source/coronaviruse/situation-reports/20200325-sitrep-65-covid-19.pdf?sfvrsn=2b74edd8_2 (accessed: Mar 26, 2020).

[12] Department of Health. 2019-nCoV acute respiratory disease, Australia Epidemiology Report 1, 2020. https://www1.health.gov.au/internet/main/publishing.nsf/Content/novel_coronavirus_2019_ncov_weekly_epidemiology_reports_australia_2020.htm (accessed: Mar 10, 2020).

[13] Department of Health. COVID-19, Australia: Epidemiology Report 2, 2020. https://www1.health.gov.au/internet/main/publishing.nsf/Content/novel_coronavirus_2019_ncov_weekly_epidemiology_reports_australia_2020.htm (accessed: Mar 10, 2020).

[14] Department of Health. COVID-19, Australia: Epidemiology Report 7, 2020. https://www1.health.gov.au/internet/main/publishing.nsf/Content/novel_coronavirus_2019_ncov_weekly_epidemiology_reports_australia_2020.htm (accessed: Mar 20, 2020).

[15] Department of Health. Coronavirus (COVID-19) current situation and case numbers, 2020. https://www.health.gov.au/news/health-alerts/novel-coronavirus-2019-ncov-health-alert/coronavirus-covid-19-current-situation-and-case-num (accessed: Mar 17, 2020).

[16] Department of Home Affairs. COVID-19 (Coronavirus) and the Australian border, 2020. https://www.homeaffairs.gov.au/news-media/current-alerts/novel-coronavirus (accessed: Mar 16, 2020).

[17] G. E. P. Box and D. R. Cox. An Analysis of Transformations. J. R. Stat. Soc. Ser. B, 1964.

[18] Denis Kwiatkowski, Peter C.B. Phillips, Peter Schmidt, and Yongcheol Shin. Testing the null hypothesis of stationarity against the alternative of a unit root. J. Econom., 1992.

[19] Rob J. Hyndman and Yeasmin Khandakar. Automatic time series forecasting: The forecast package for R. J. Stat. Softw., 2008.

[20] Peter Caley, Niels G. Becker,and David J. Philp. The Waiting Time for Inter-Country Spread of Pandemic Influenza. PLoS One, 2(1):e143, 2007.

[21] C R MacIntyre. On a knife’s edge of a COVID-19 pandemic: is containment still possible? Public Heal. Res. Pract., 30:3012000, 2020.

